# Capsule-independent bacteriophages reveal unexpected diversity of Salmonella Typhi phage ecology

**DOI:** 10.64898/2026.08.19.26360815

**Authors:** Shantu Dey, Shuborno Islam, Al Amin, Mohammad Daniyal Karim, Sadnane Hussain Pranto, Rathindranath Kabiraj, Neoyman Nasir, Hakka Naziat, Arif M Tanmoy, Samir K Saha, Senjuti Saha, Yogesh Hooda

## Abstract

Bacteriophages that infect *Salmonella enterica* serovar Typhi (*S.* Typhi), the cause of typhoid fever, are regarded as specialized, because all previously characterized phages depend on the Vi capsular polysaccharide for infection. Whether capsule-independent infection strategies exist has remained unclear. Here we identify environmental phages that infect *S.* Typhi both in the presence and absence of Vi. Screening 140 urban wastewater samples from Dhaka, Bangladesh, where typhoid is endemic, we recovered phages infecting a Vi-deficient *S.* Typhi strain from 41 samples (29%). All 41 phages also infected the isogenic Vi-expressing host, although 28 did so with 10- to 10⁵-fold lower infection efficiency, and suppressing capsule expression increased susceptibility to 23, indicating an inhibitory effect of Vi on infection by many these phages. All 41 phages infected *S.* Paratyphi A and nine infected a monophasic *S.* Typhimurium, a broader host range than the Vi-dependent phages, which were restricted to Vi-expressing Typhi. Across 26 circulating genotypes, capsule suppression increased susceptible genotypes per phage by 1.51 on average (Wilcoxon *p* = 5.76 × 10⁻⁶), though four genotypes remained resistant to all phages tested, indicating additional determinants of susceptibility. Whole-genome sequencing of 27 phages identified three genera in two families, predominantly *Teetrevirus* (19/27); TerL phylogeny separated these from classical Vi-dependent phage lineages. Together, these findings reveal a broader-host-range component of Typhi phage ecology and show that Vi dependence is not a universal feature of phages capable of infecting *S.* Typhi.

**Importance:** The Vi capsule is a defining surface antigen of *Salmonella* Typhi and is required for infection by characterized Typhi phages. Here, we show that this dependence is not universal. In a typhoid-endemic setting, we identified environmental phages capable of infecting *S.* Typhi in the absence of Vi, revealing a previously underrecognized infection strategy. These Vi-independent phages showed broader host ranges than classical Vi-dependent phages, with susceptibility varying across *S.* Typhi lineages. Genomic characterization identified that these phages were from lineages distinct from Vi-dependent phages. Together, these findings reveal greater diversity in *S.* Typhi-phage interactions than previously appreciated and suggest that Vi-dependent specialists represent only one component of a broader Typhi-phage ecological landscape. As these phages were identified before the national introduction of the Vi-typhoid conjugate vaccine, they also provide a baseline for future studies examining whether population-level targeting of the Vi capsule is accompanied by changes in Typhi-phage ecology.

## Introduction

Bacteriophages that infect *Salmonella* Typhi (*S.* Typhi), the causative agent of typhoid fever, have been studied since the 1940s, initially to differentiate and track strains during typhoid fever outbreaks (1, 2). This large body of work established the classical Vi phage-typing scheme, which differentiated *S.* Typhi strains according to their susceptibility to Vi phages (3, 4). More recently, several groups have isolated and genomically characterized Typhi phages from environmental water in typhoid-endemic regions, showing that Typhi phages are abundant and stable in the environment (5, 6). Historical Typhi phages have since been assigned to several genera, three of which, *Kayfunavirus*, *Teseptimavirus*, and *Macdonaldcampvirus*, have also been recovered recently in Nepal and Bangladesh (6–9). Despite their genomic diversity, these classically characterized Typhi phages share a striking feature: infection depends on the Vi capsular polysaccharide. This strict reliance on the Vi capsule reflects a high degree of host specificity and places the characterized Typhi phages among specialist phages with a restricted host range. In other bacterial systems, such as *Klebsiella*, phages can instead act as generalists, infecting multiple hosts or strains (10). Whether such generalist strategies also exist among *S.* Typhi phages has remained largely unexplored.

The Vi capsule is encoded by the *viaB* locus on Salmonella Pathogenicity Island 7 and forms a surface-exposed polysaccharide layer around *S.* Typhi (11). Its position at the cell surface makes it readily accessible to bacteriophages. Biochemical studies show that Vi-dependent phages carry acetyl esterase domains that recognize the O-acetylated Vi polymer and enzymatically modify it to initiate infection (9, 12). Deletion of the Vi biosynthesis locus abolishes susceptibility to these phages, indicating that the interaction with the Vi capsule is essential for their infection (8).

Several lines of evidence, however, suggest that this dependence may not be universal and that alternative, potentially generalist strategies could exist. First, the transcription of the Vi capsule is regulated by environmental signals, including osmolarity and temperature: high osmolarity and lower temperature repress *tviA*, a regulatory gene of the *viaB* locus, and reduce capsule production, whereas host-associated conditions promote Vi synthesis (13, 14). Second, naturally occurring Vi-negative *S.* Typhi strains have been reported in multiple settings, including Pakistan (15, 16) and Malaysia (17), showing that *S.* Typhi strain lacking the Vi capsule do circulate. Finally, the flagellotropic phage YSD1, isolated from a river in Cambridge, UK, infects *S.* Typhi independently of the Vi capsule; although not Typhi-specific and able to infect multiple *Salmonella* serovars, its existence shows that non-Vi receptor pathways can mediate infection (18). However, YSD1 has remained an isolated example. Whether Vi-independent infection represents a rare exception or a broader and diverse feature of phages capable of infecting *S.* Typhi is unknown. This question has gained additional relevance with the introduction of Vi-targeted typhoid conjugate vaccines, which creates an opportunity to examine whether population-level targeting of the Vi capsule is accompanied by changes in *S.* Typhi–phage ecology (19, 20).

Building on these observations, we hypothesized that diverse Vi-independent Typhi phages capable of infecting *S.* Typhi circulate in endemic environments. We therefore collected wastewater from an area of Dhaka, Bangladesh, with sustained typhoid transmission and enriched for phages capable of infecting *S.* Typhi in the absence of the Vi capsule. We then examined how Vi expression influences phage infectivity and host range across *S.* Typhi isolates and other *Salmonella* serovars and characterized the genomic diversity and evolutionary relationships of the recovered phages. Together, these experiments were designed to determine whether Vi-independent infection represents an isolated exception or a broader feature of phages capable of infecting *S.* Typhi.

## Methodology

### Bacterial strains and Vi capsule states

Two isogenic *S.* Typhi strains were used as primary hosts throughout. BRD948 is an attenuated laboratory strain that expresses the Vi capsule (hereafter Vi⁺). BA256 is a Vi-deficient derivative (hereafter ΔVi) carrying a kanamycin cassette disrupting the *tviB* gene within the *viaB* locus (8). A third, conditional state, BRD948 grown under capsule-suppressing conditions (hereafter ViO; LB with 300 mM NaCl, 23 °C) (21), was used to reduce Vi expression without altering the genotype. Standard (Vi-expressing) conditions were LB with 171 mM NaCl at 37 °C. Three operational terms are used throughout. Infectivity is the ability of a phage to form detectable plaques on a given host (a qualitative, yes/no property). Plating efficiency is how readily this occurs, quantified as the minimum phage titre (PFU) required to produce plaques on that host (higher efficiency means plaques form at lower titres). Host range is the breadth of bacterial strains, serovars, and species that a phage can productively infect.

### Environmental wastewater sampling

A total of 140 wastewater samples from open drains were collected between 12^th^ and 20^th^ of July 2023 from an area of Dhaka, Bangladesh, with known typhoid transmission. Sites were chosen to avoid cross-contamination and repeated collection, maintaining a mean inter-site distance of 0.44 km (mean *k*-nearest-neighbour distance, *k* = 3). Ten mL of wastewater was collected from each of 140 drainage points using a sterile 15-mL conical tube attached to a rope. Samples were transferred into fresh sterile tubes using procedures designed to prevent cross-contamination between sites. and transported to the laboratory within 24 hours. On arrival, samples were centrifuged at 500 × g for 10 min to pellet large debris, and the supernatant was filtered through a 0.22 µm polyethersulfone (PES) syringe filter. Filtered samples were stored at 4 °C until processed within 48 hours.

### Phage isolation and purification

Phages were isolated using the double-layer agar (DLA) method as described previously (5). Briefly, 500 µL of filtered sample was mixed with 450 µL of Luria–Bertani (LB) broth and 50 µL of overnight liquid culture of *S.* Typhi (Vi⁺ or ΔVi, separately) and incubated at 37 °C for 2 h. To lyse remaining bacteria, 2–3 drops of chloroform (Carl Roth, USA) were added; the mixture was vortexed, incubated for 10 min at room temperature, and centrifuged at 10,000 × g for 10 min. Approximately 800 µL of phage-containing supernatant was transferred to a fresh tube.

For detection, 100 µL of enriched supernatant was mixed with 200 µL of overnight culture of each host and incubated for 20 min at room temperature to allow adsorption (22). The mixture was added to 4 mL of molten soft agar (0.7% LB agar) and overlaid on hard agar plates (1.5% LB agar). Plates were incubated at 37 °C for 16–18 h and examined for plaques: on ΔVi for Vi-independent phages and on Vi⁺ for Vi-dependent phages. Individual plaques of distinct morphology were picked with sterile tips and resuspended in 150 µL of LB broth. Following chloroform treatment and centrifugation, 100 µL of supernatant, a clonal stock of a single phage, was retained. Phage titres were determined by spotting 2 µL of 100-fold serial dilutions onto bacterial lawns.

#### Suppression and confirmation of Vi expression *in vitro*

To assess capsule-dependent infection, Vi⁺ was cultured under capsule-suppressing conditions (ViO; LB with 300 mM NaCl, 23 °C). Vi-independent phage suspensions (2 µL; 10⁷–10⁰ PFU/mL) were spotted onto lawns of Vi⁺, ΔVi, and ViO. Previously characterized and sequenced Typhi-specific phages of three genera (*Kayfunavirus*, *Teseptimavirus*, and *Macdonaldcampvirus*) were included as Vi-dependent reference phages (7). Plates of Vi⁺ and ΔVi were incubated at 37 °C for 16–18 h; plates of ViO were incubated at 23 °C for the same duration.

Vi expression was confirmed by slide agglutination using *S.* Typhi Vi antiserum. A single colony from each condition was inoculated into liquid LB and incubated overnight under either standard (Vi-expressing) or capsule-suppressing conditions; the corresponding salt and temperature were maintained throughout, and cultures were streaked onto LB agar of matching salt concentration and incubated at the matching temperature to obtain isolated colonies. A fresh single colony from each condition was resuspended in 20 µL of normal saline on a glass slide, mixed with 10 µL of Vi antiserum (Deben Diagnostics Ltd, UK; Ref 294470), and scored for visible agglutination within 1–2 min. Three states were tested in parallel, Vi⁺, ViO, and ΔVi, and images were captured on a ChemiDoc imaging system.

### Host range and genotype infectivity

For host-range analysis, 2 µL of 10⁷ PFU/mL phage suspension was spotted onto lawns of *S. enterica* serovars Paratyphi A, Worthington, Virchow, Enteritidis, Typhimurium LT2, and monophasic variant of *S. enterica* serovar Typhimurium 1,4,[5],12:i:-, and onto non-*Salmonella* Gram-negative species: *Escherichia coli*, *Pseudomonas aeruginosa*., *Klebsiella pneumoniae*, *Enterobacter cloacae*, and *Citrobacter freundii*. After incubation at 37 °C for 16–18 h, plaque formation or zones of clearing were scored.

For genotype infectivity, Vi-independent phages were tested against 26 *S.* Typhi genotypes circulating in Bangladesh. Isolates were grown under both standard (Vi-expressing) and capsule-suppressing conditions. Phage suspensions were spotted onto each lawn and incubated under the corresponding condition, and plaque formation was scored. Vi expression in all isolates under both conditions was verified with Vi antiserum.

### Whole-genome sequencing of Vi-independent phages

Vi-independent phages were sequenced as described previously (23). Briefly, phages were propagated to confluent lysis by the DLA method. Plates were flooded with 4 mL of LB and incubated at 4 °C for 5 h to elute phage particles. The eluate was transferred to a fresh tube, treated with chloroform, vortexed, and centrifuged. The supernatant was transferred to a fresh tube and treated with 1 µL of DNase (1 U/µL), 50 µL of DNA Digestion Buffer and 0.5 µL of RNase A (20 mg/mL) to remove bacterial nucleic acid. Enzymes were inactivated with 20 µL of 0.5 M EDTA, followed by 1.25 µL of Proteinase K (20 mg/mL) to digest capsid protein. Genomic DNA was purified with the QIAamp DNA Mini Kit (Qiagen, Germany; Cat 51306). Libraries were prepared with the NEBNext Ultra II FS DNA Library Prep Kit (New England Biolabs, USA; E7805L) and sequenced on an Illumina platform (NextSeq 2000 or iSeq 100) to generate 150 bp paired-end reads.

### Bioinformatic analysis

Raw FASTQ files were quality-assessed with FastQC (v0.11.5); Trimmomatic (v0.39) removed adapters and low-quality reads (Phred < 20). Reads were assembled into a single contig per phage with Unicycler (v0.5.1). Functional annotation used Pharokka (v1.8.2), and taxonomic classification used Kraken2 (v2.17.0) on the single circular contig of each phage.

For phylogenetics, amino-acid sequences of the terminase large subunit (TerL), a widely used conserved marker for dsDNA phage phylogeny (24), were extracted from all phages sequenced here and from previously sequenced Typhi phages (6–9, 18). The comparison set comprised 47 previously sequenced Vi-dependent phages from historical collections and environmental studies in Bangladesh and Nepal, together with the flagellotropic phage YSD1. TerL sequences were aligned with MAFFT (v7.505), and a tree was inferred in IQ-TREE (v3.0.1) with ModelFinder, 1000 ultrafast-bootstrap replicates, and the SH-aLRT test. The tree was mid-point rooted and visualized in iTOL (v7.5).

### Data visualization and statistical analysis

Analyses were performed in R (v4.5.2). Data were handled with *dplyr*, *tidyr*, and *reshape2*. For plaque-assay data (Fig. 2), qualitative outcomes were converted to semi-quantitative values (“clear” = 80, “TNTC” = 50, “no plaque” = 0) and log₁₀-transformed [log₁₀(PFU + 1)] for display; heatmaps and plots used *ggplot2*, and multi-panel figures used *patchwork*. Genotype infectivity across 26 genotypes was visualized as binary heatmaps (plaque/no plaque); susceptibility was defined as plaque formation at 10⁷ PFU/mL. Clustering (Fig. 3) used Jaccard distance (*vegan*) followed by hierarchical clustering with optimal leaf ordering (*seriation*); heatmaps used *ComplexHeatmap* with colour functions from *circlize*. For host range (Fig. 4), binary infectivity data were displayed without clustering using *gplots* (heatmap.2); phages were ordered by infectivity pattern, and genus annotations used *RColorBrewer*. Paired comparisons between Vi-expressing and capsule-suppressing conditions used a two-sided Wilcoxon signed-rank test.

**Figure 1:**
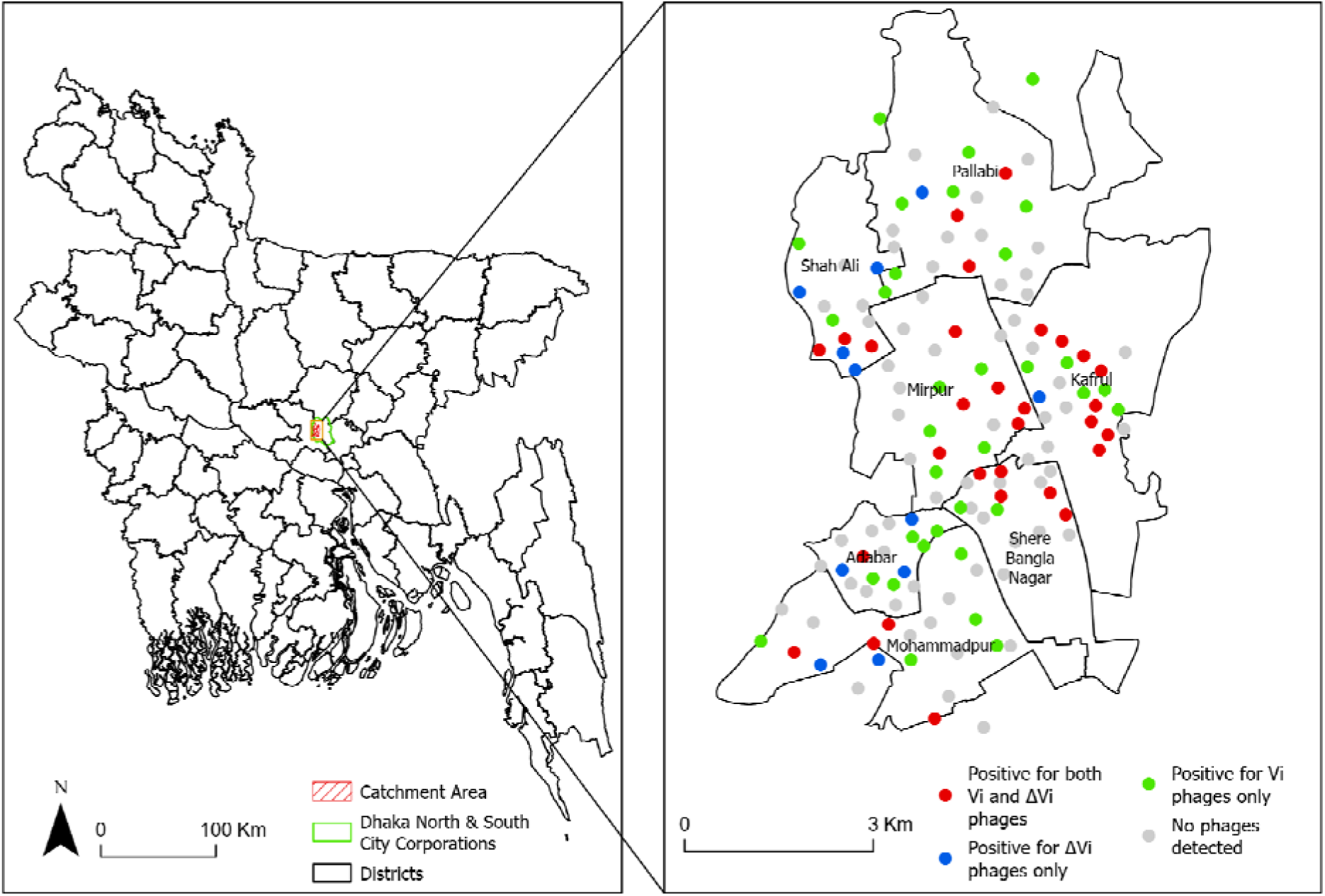
Geographical distribution of wastewater sampling sites and detection of Vi-dependent and Vi-independent *S.* Typhi phages in Dhaka, Bangladesh. Each point is a wastewater sampling site (n = 140). Colors indicate detection outcomes: red, sample positive for both Vi-independent and Vi-dependent phages; green, Vi-dependent only; blue, Vi-independent only; grey, no detectable phages. Sampling locations were assigned on a manually generated map (2023, Google Earth) and visualized in ArcGIS; when overlaid on an updated digital map (2025), a few points fall outside the catchment boundaries due to differences between map versions. (Map source: Bangladesh Subnational Administrative Boundaries, https://data.humdata.org/dataset/cod-ab-bgd.)

**Figure 2.**
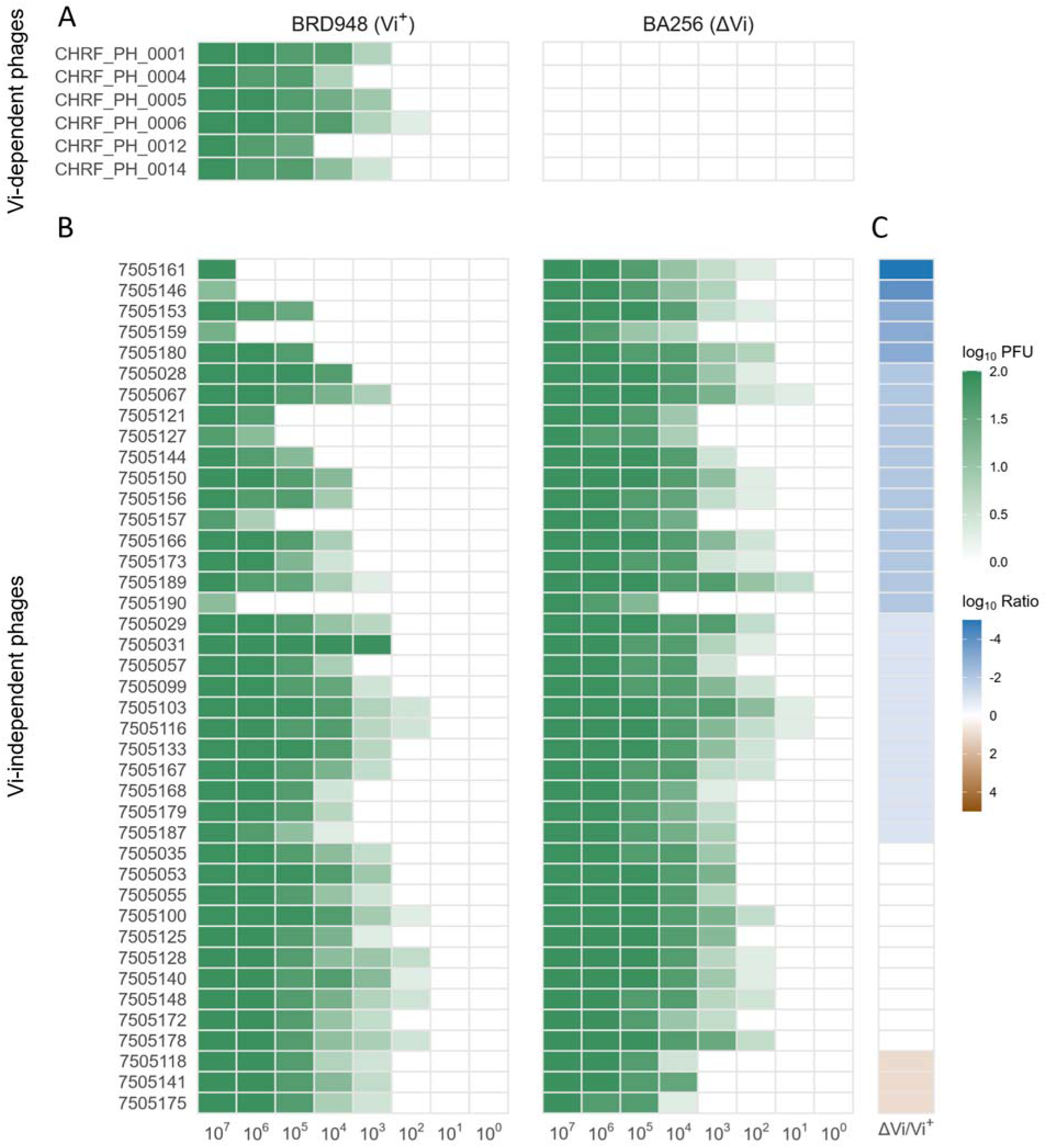
Capsule-dependent modulation of Vi-independent phage infection efficiency. (A, **B)** Infectivity heatmaps on Vi-expressing (Vi⁺, BRD948) and Vi-deficient (ΔVi, BA256) hosts, shown as log₁₀(PFU + 1) for 6 Vi-dependent phages **(A)** and 41 Vi-independent phages **(B)**. Columns are 10-fold serial dilutions of lysate (10⁷–10⁰ PFU/mL); rows are individual phages. Darker green indicates plaque formation at lower dilution (higher efficiency); lighter colors indicate reduced or absent plaque formation. **(C)** Host-preference index. For each phage, the lowest PFU producing plaques on ΔVi was compared with Vi⁺ and expressed as log₁₀(ΔVi/Vi⁺). Blue (−5 to −1), higher efficiency on ΔVi; white, similar on both; light brown (> 0), higher efficiency on Vi⁺. Generated in R (v4.5.2) with *ggplot2*.

**Figure 3.**
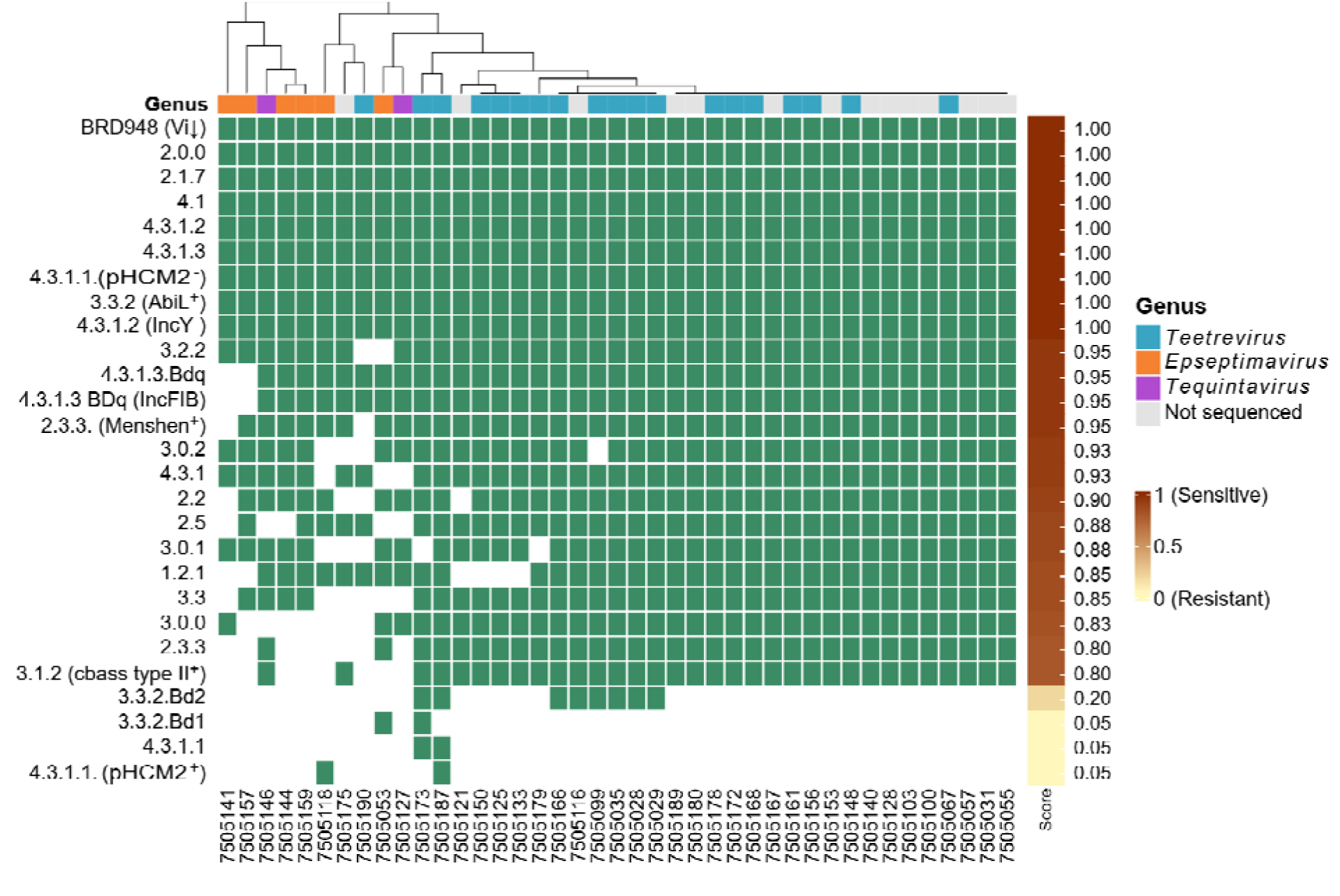
Host range of Vi-independent phages across circulating S. Typhi genotypes under capsule-suppressing conditions. Infectivity of 41 Vi-independent phages against 26 circulating S. Typhi genotypes grown under capsule-suppressing conditions (LB, 300 mM NaCl, 23 °C). Each phage was spotted at 10⁷ PFU/mL, and infectivity was scored by presence/absence of plaques. Green, detectable killing (sensitive); white, resistant. Row are genotypes; columns are phages. The adjacent brown–yellow strip shows an infectivity score (0–1) relative to the BRD948 reference, which showed the highest susceptibility; scores near 1 resemble BRD948, scores near 0 indicate resistance. Hierarchical clustering highlights lineage-specific variation. The colored bar above the heatmap indicates phage genus. Generated in R (v4.5.2) with ComplexHeatmap; clustering used Jaccard distance (vegan).

**Figure 4.**
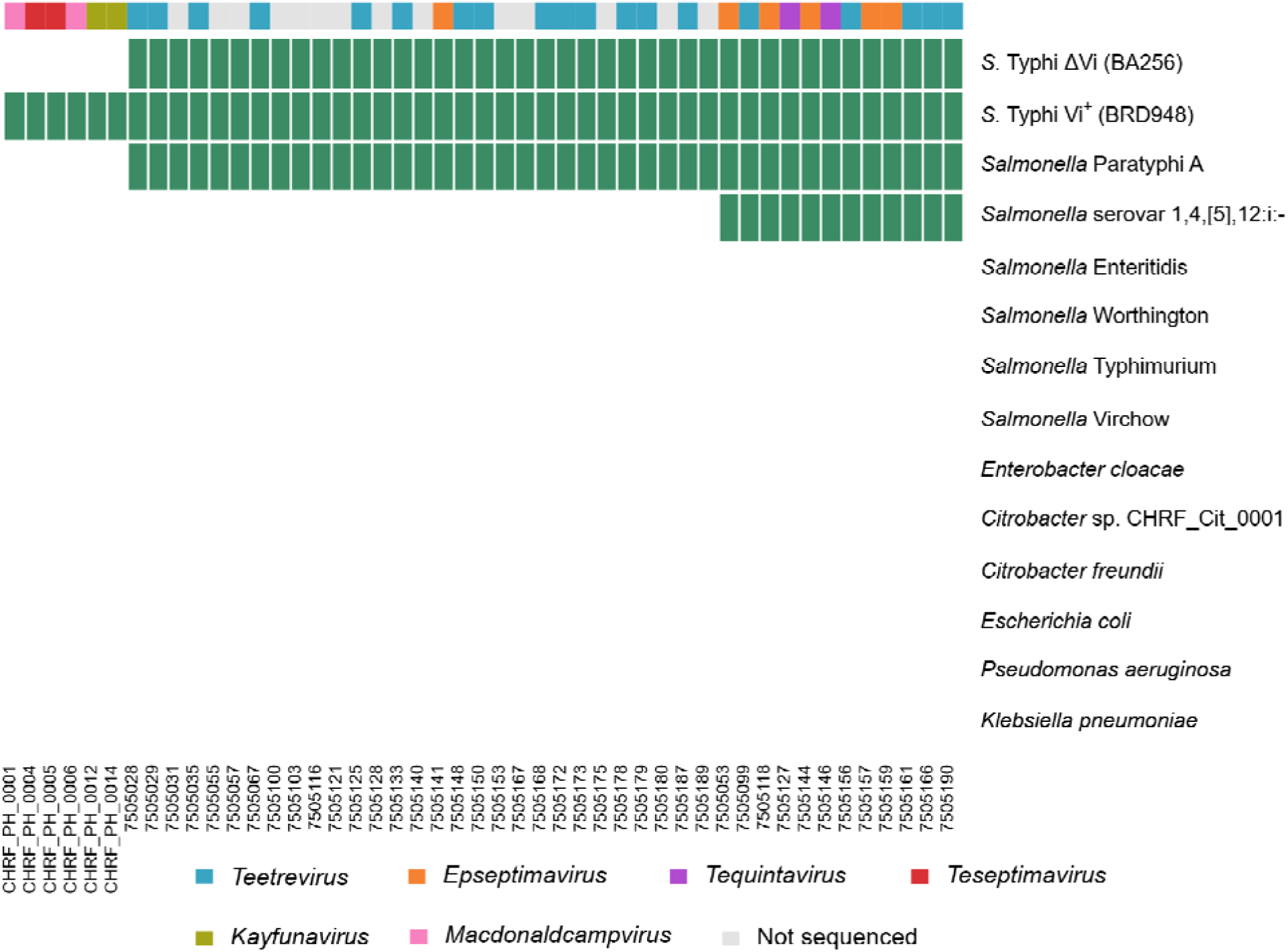
Host specificity of Vi-dependent and Vi-independent phages across Salmonella and non-Salmonella species. Infectivity of 6 Vi-dependent and 41 Vi-independent phages against a panel of Salmonella serovars and non-Salmonella Gram-negative species. Each phage was spotted at 10⁷ PFU/mL and scored by plaque formation. Green, detectable lytic activity; white, no infection. Rows are species/serovars; columns are phages. The colored bar above the heatmap indicates phage genus. Generated in R (v4.5.2) with gplots (heatmap.2).

## Results

### Phages capable of infecting Vi-deficient S. Typhi are widely detected in urban wastewater

To determine whether phages capable of infecting *S.* Typhi independently of the capsule can be recovered from the environment, we screened 140 wastewater samples from an of area of Dhaka with documented high typhoid incidence (25). Of these, 41 of 140 samples (29%) yielded plaques on the Vi-deficient host (ΔVi, BA256), demonstrating that phages capable of infecting *S.* Typhi independently of Vi were detectable at a substantial fraction of sampled sites. One phage isolate was purified from each positive sample, yielding 41 isolates. In parallel, screening on the Vi-expressing host (Vi⁺, BRD948) detected phages in 62 samples (44%; **Fig. 1**), consistent with the expected abundance of Vi-dependent phages (5) (3). Vi-independent phages therefore co-occur with classical Vi-dependent phages in this setting.

### The Vi capsule limits infection by Vi-independent phages

To determine how the Vi expression influences phage susceptibility, we compared the infectivity of 47 phages, capable of infecting *S.* Typhi across the three Vi states (Vi⁺, ΔVi, ViO): 41 Vi-independent phages isolated here (**Fig. 2B**) and 6 previously characterized Vi-dependent reference phages representing *Kayfunavirus*, *Teseptimavirus* and *Macdonaldcampvirus* (**Fig. 2A**)(7). Vi antiserum agglutination confirmed Vi expression under standard conditions and loss of detectable Vi reactivity in ΔVi and under capsule-suppressing conditions (**Supplementary Fig. S1A**).

The Vi-dependent reference phages formed plaques only on Vi^+^ cells (**Fig. 2A–B; Supplementary Fig. S1B–C**). In contrast, all 41 Vi-independent phages infected both ΔVi and Vi^+^ hosts, although their relative plating efficiencies differed substantially (**Fig. 2A–C**). To compare relative plating efficiency between the two hosts, we calculated the ratio of the minimum phage concentrations required to produce plaques on ΔVi and Vi^+^. For 28 of 41 phages (68%), detectable plaque formation required a 10- to 10⁵-fold lower phage concentration on ΔVi than on Vi^+^. Ten of 41 (24%) were similar on both hosts (ratio ≈ 0); and 3 of 41 (7%) formed plaques at lower titres on Vi⁺, i.e. a modest preference for the encapsulated host (ratio > 0).

Consistent with this pattern, growth under Vi-suppressing conditions increased susceptibility to 23 of 41 phages (56%) relative to standard Vi-expressing conditions; 11 phages (27%) showed no difference and seven (17%) showed greater activity under Vi-expressing conditions (**Supplementary Fig. S1B–D**). These observations indicate that Vi expression limits infection by most of the Vi-independent phages tested.

#### Capsule-suppressing conditions broaden across *S.* Typhi genotypes

To assess the breadth of Vi-independent phage activity, we tested the 41 phages against 26 *S.* Typhi genotypes circulating in Bangladesh under both standard (Vi-expressing) and capsule-suppressing conditions. Under standard conditions, more than 20 of the 26 genotypes were susceptible to Vi-independent phages (**Supplementary Fig. S2**); genotypes 1.2.1, 3.3.2.Bd1, 3.3.2.Bd2, and 4.3.1.1 were notably resistant. Within the 4.3.1.1 lineage, the pHCM2⁺ isolate was resistant whereas the pHCM2⁻ isolate was susceptible; pHCM2 is a phage-plasmid carrying genes for an intact prophage (26). Under capsule-suppressing conditions, susceptibility increased across multiple genotypes (**Fig. 3**): genotype 1.2.1 shifted from minimal to detectable susceptibility, with similar increases in 3.3.2.Bd2 and several 4.3.1 lineages.

Relative infectivity scores (0–1, normalized to the Vi⁺ reference strain BRD948) were higher under capsule-suppressing conditions across ∼20 genotypes (**Fig. 3, Supplementary Fig. S2**). Hierarchical clustering of the genotype-by-phage susceptibility profiles identified 17 clusters, reflecting functional diversity within the Vi-independent population (**Fig. 3, Supplementary Fig. S2**). To quantify the effect of capsule suppression, we compared, for each phage, the number of susceptible genotypes under the two conditions (**Supplementary Fig. S3A–B**): each phage infected ∼20 genotypes unde standard conditions and ∼22 under capsule-suppressing conditions, and most phages fell above the line of equality (**Supplementary Fig. S3B**). The number of susceptible genotype per phage increased by 1.51 on average (7.84%; two-sided Wilcoxon p = 5.76 × 10⁻⁶). Vi-independent phages thus infect a broad range of circulating S. Typhi genotypes, and capsule suppression widens that range further.

#### Vi-independent phages show broader activity across *Salmonella* serovars

All 47 phages were tested against a panel of *Salmonella* serovars and non-*Salmonella* Gram-negative bacteria to compare host range (**Fig. 4**). The six Vi-dependent reference phages formed plaques only on Vi-expressing *S.* Typhi and showed no detectable activity against ΔVi *S.* Typhi or the other bacterial strains tested. In contrast, all 41 Vi-independent phages formed plaques on both *S.* Typhi host states and on the tested *S.* Paratyphi A, and 9 of 41 also infected *S. enterica* subspecies I serovar 1,4,[5],12:i:-. None of the phages infected *S.* Enteritidis, *S.* Worthington, *S.* Typhimurium, or *S.* Virchow, nor the non-*Salmonella* species *Enterobacter cloacae*, *Citrobacter freundii*, *Escherichia coli*, *Pseudomonas aeruginosa*, or *Klebsiella pneumoniae*. Vi-dependent phages therefore appear to specialize in infecting for Vi-expressing *S.* Typhi, whereas infectivity of Vi-independent phages extend to other *Salmonella* serovars, likely by targeting surface receptors shared among *S.* Typhi, *S.* Paratyphi A, and monophasic variant of *S. enterica* serovar Typhimurium 1,4,[5],12:i:-.

#### Vi-independent phages form clades distinct from Vi-dependent phages

To place the Vi-independent phages relative to previously studied Typhi phages, we sequenced 27 of them, chosen to represent the susceptibility clusters in Fig. 3. Taxonomic classification assigned them to three genera across two families (**Supplementary Table 1**). Nineteen belonged to *Teetrevirus* (family *Autographiviridae*), with genomes of 37–39 kb encoding 52–55 open reading frames (ORFs). The remaining phages belonged to *Epseptimavirus* (six phages) and *Tequintavirus* (two phages), both in the family *Demerecviridae*, with larger genomes of 111–115 kb encoding 202–212 ORFs; most ORFs had no annotated function. *Autographiviridae* members carried a single-subunit RNA polymerase gene (27).

We constructed a phylogeny from TerL amino-acid sequences of 75 phages: the 27 Vi-independent phages sequenced here, 47 previously studied Vi-dependent phages {1930s– 1950s isolates (8, 9), and isolates from Bangladesh (7) and Nepal (6), and the flagellotropic phage YSD1 (18). The Vi-independent phages occupied clades distinct from the classical Vi-dependent phages, demonstrating that Vi-independent infection occurs acros phylogenetically distinct phage lineages (**Fig. 5**). Phages clustered primarily by genus, with the Vi-independent Teetrevirus phages forming a clade distinct from the Vi-dependent Teseptimavirus and Kayfunavirus references. The Vi-independent Demerecviridae phages (Epseptimavirus, Tequintavirus) formed a cluster with partial intermixing, alongside the Chivirus YSD1.

**Figure 5:**
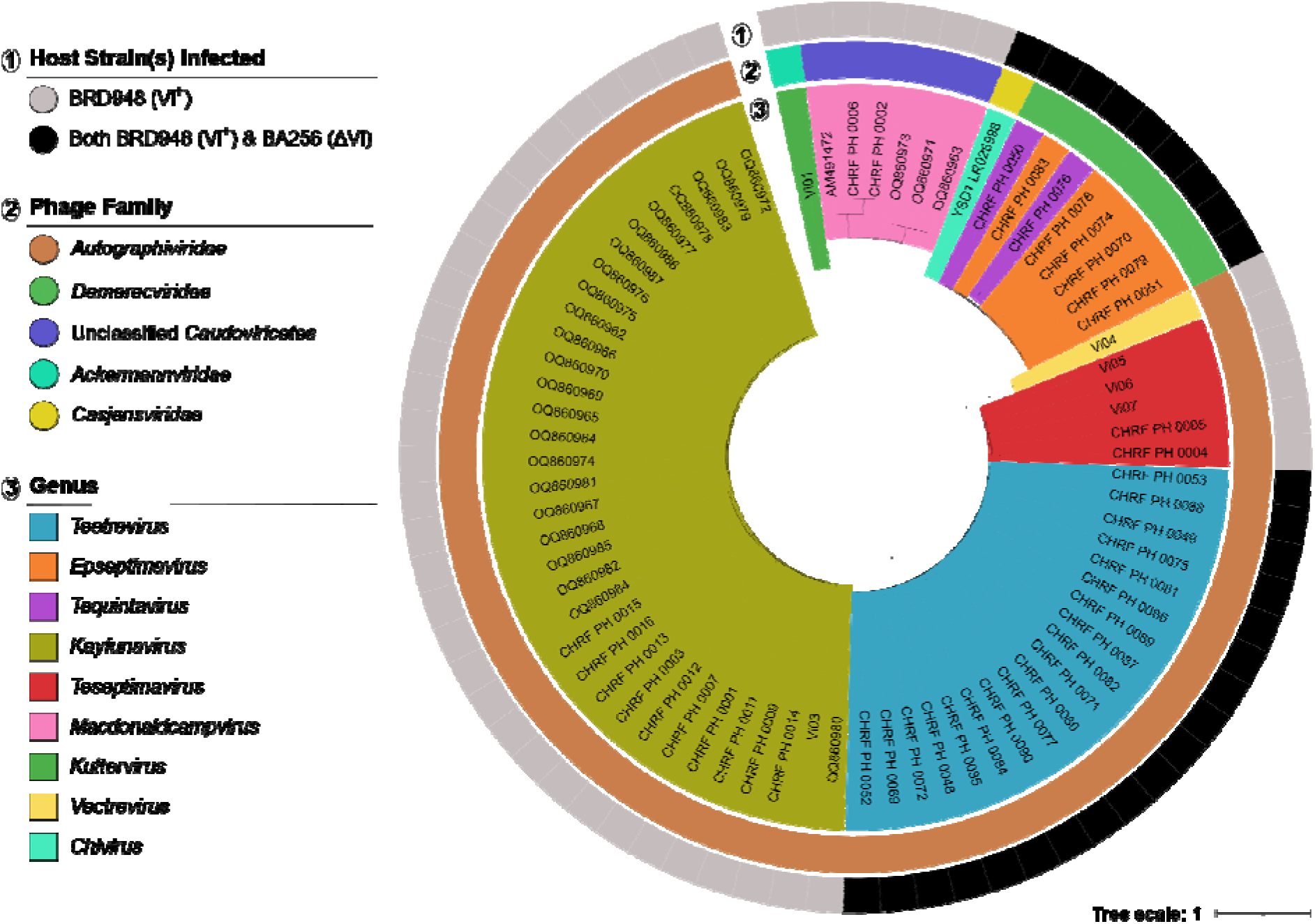
Phylogenetic relationship among Vi-dependent and Vi-independent S. Typhi phages based on the terminase large subunit (TerL). Rooted phylogeny of TerL amino-acid sequences from 75 Typhi phages (27 Vi-independent phages sequenced here, 47 previously studied Vi-dependent phages, and 1 flagellotropic phage). Colour strips indicate the host strain(s) infected and the phage family; phage genera are marked a colour-labelled ranges. Sample IDs for the 27 Vi-independent phages sequenced here are listed in **Supplementary Table 1**.

Genome organization also differed substantially between the two Vi-independent phage families. *Teetrevirus* genomes were smaller (37–39 kb) and showed highly conserved gene order, with discrete replication and structural modules and localized accessory regions. In contrast, the larger *Demerecviridae* genomes (111–115 kb) contained extended replication and structural regions, more than 20 tRNA genes, and greater variation in accessory gene content (**Supplementary Figs. S4–S5**). These differences further demonstrate that Vi-independent infection is not restricted to a single genomic architecture.

## Discussion

We identify and characterize a previously underrecognized group of bacteriophages capable of infecting *S.* Typhi independently of the Vi capsule. Classically characterized Typhi phages depend on Vi for infection, establishing Vi recognition as a central paradigm in Typhi-phage biology (9, 28). In this study, recovery of Vi-independent phages from 41 of 140 wastewater samples shows that this alternative infection strategy is not limited to an isolated phage or sampling site but is widely detectable in the sampled environment. Overall, our findings reveal at least two distinct modes of phage interaction with *S.* Typhi: classical Vi-dependent specialization and Vi-independent infection associated with a broader host range. These findings reshape the understanding of Typhi–phage interactions in two ways. First, the Vi capsule is not the sole determinant of susceptibility. Our findings show that Vi is not universally required for phage infection of *S.* Typhi and, for many Vi-independent phages, its presence is associated with reduced plaque formation. For most of these phages, detectable plaques formed at substantially lower phage concentrations on the isogenic Vi-deficient host than on the Vi-expressing host, with a similar trend observed under Vi-suppressing growth conditions. One possible explanation is that the Vi capsule physically limits access to underlying surface receptors used by these phages. Candidate receptors could include conserved outer-membrane structures such as lipopolysaccharide or outer-membrane proteins, although direct identification of these receptors will require genetic and biochemical validation (29, 30).

A second distinction lies in host range. The Vi-dependent reference phages were restricted to Vi-expressing *S.* Typhi, whereas the Vi-independent phages showed activity against additional *Salmonella* hosts, most consistently the tested *S.* Paratyphi A strain. Their host range was nevertheless structured rather than unrestricted, as activity was not observed across several other *Salmonella* serovars. These patterns are consistent with a specialist– generalist continuum (10), with classical Vi-dependent phages occupying the more specialized end and Vi-independent phages showing broader host use. How Vi dependence evolved, whether through acquisition of Vi-specific adaptations or loss of broader receptor usage, remains an open question for future comparative genomic and functional studies.

The historical predominance of Vi-dependent phages may partly reflect the methods used for their isolation and characterization. Vi-dependent phages may dominate enrichment cultures because the Vi capsule is highly expressed under host-like conditions and presents an abundant, exposed receptor that supports rapid adsorption and replication; in mixed populations, such phages may outcompete capsule-independent phages during laboratory enrichment, biasing detection. This may also explain why phage-based environmental surveillance for *S.* Typhi has been so effective (5, 6): the Vi capsule is a distinctive, relatively conserved surface feature, allowing Vi-dependent phages to serve as sensitive indicators of *S.* Typhi.

Genomic analysis supports an evolutionary distinction between the two groups. The TerL phylogeny resolved Vi-independent phages into clades separate from classical Vi-dependent lineages, indicating independent evolutionary trajectories, and the Vi-independent phages spanned two families (*Autographiviridae* and *Demerecviridae*), showing that capsule independence is not confined to a single genomic background. Host range varied even within a family: within *Autographiviridae*, closely related phages showed either narrow (Vi-dependent) or broad (Vi-independent) infectivity, also suggesting a continuum of specialization rather than strict taxonomic segregation. Differences in genome size, and gene content suggest divergent infection strategies, potentially linked to alternative receptor usage; functional validation is needed to confirm the roles of these regions (31, 32).

Vi expression was also not sufficient to explain the full variation in susceptibility among *S.* Typhi isolates. Despite this broad host range, selected isolates of four *S.* Typhi genotypes remained resistant even under capsule-suppressing conditions. The contrasting susceptibility profiles observed among isolates belonging to the same 4.3.1.1 genotype, likely caused by the presence of cryptic phage-plasmid pHCM2 (26), further suggest that strain-specific features contribute to this variation. Possible determinants include differences in the underlying phage receptor, variation in lipopolysaccharide or other surface structures, and intracellular anti-phage defence systems. Identifying these additional determinants will be important for understanding the molecular basis of Vi-independent infection.

The findings of our study must be considered within the context of several limitations. First, the plating-efficiency measurements used here reflect the minimum phage concentration required for detectable plaque formation under the conditions tested and do not directly measure adsorption efficiency, efficiency of plating, burst size, or replication kinetics (33–35). These additional measurements will be important for defining how the Vi capsule influences individual stages of infection. Second, although the isogenic Vi-deficient strain provides direct evidence that capsule state influences susceptibility, the Vi-suppressing growth conditions also differ in temperature and osmolarity and may alter other bacterial surface features; these experiments should therefore be interpreted as supportive rather than as an isolated effect of Vi suppression. Third, host-range and strain-level susceptibility were assessed using a defined bacterial panel, and susceptibility of individual isolates should not be assumed to represent all members of a genotype or serovar. Finally, the receptors used by the Vi-independent phages remain unknown, and the proposed roles of underlying surface structures such as lipopolysaccharide or outer-membrane proteins will require genetic and biochemical validation.

Beyond revealing previously underappreciated diversity in *S.* Typhi–phage interactions, these findings also provide a basis for asking how this ecology may change when selective pressures on the Vi capsule change at the population level. This question is particularly timely in Bangladesh, where a Vi-targeted typhoid conjugate vaccine was introduced nationally after the wastewater sampling conducted for this study (36, 37). Because our samples were collected before vaccine introduction, they provide a pre-vaccine snapshot of Vi-independent phage diversity against which future observations can be compared. Whether widespread anti-Vi immunity will alter Vi expression or the frequency of Vi-low or Vi-negative *S.* Typhi, and whether such changes would in turn affect the relative abundance or activity of Vi-dependent and Vi-independent phages, remains unknown. Longitudinal sampling of environmental phages together with phenotypic characterization of contemporaneous clinical isolates would provide a direct way to test these possibilities.

Together, the observations made in this study reveal a more diverse landscape of *S.* Typhi– phage interactions than previously recognized, in which capsule state is one of several factors shaping susceptibility of *S.* Typhi to its phages. Future studies defining the receptors and bacterial determinants underlying Vi-independent infection will be important for understanding how these distinct phage infection strategies arise and are maintained in natural environments.

## Data Availability

Raw data for the 27 Typhi phages sequenced in this study are available on NCBI under BioProject PRJNA1081195 (SRA accessions SAMN57248883–SAMN57248909). Sequences of the 26 phages from Nepal and 14 phages from Bangladesh are available under BioProjects PRJNA933946 (accessions OQ860962–87) and PRJNA1081195 (accessions PQ336901–14), respectively. The YSD1 phage sequence is available on NCBI (accession LR026998). Sequences of the 7 phages from the USA, Canada, and Germany are available on NCBI (accession AM491472) and via Sanger FTP (IDs Vi01, Vi03–Vi07; https://ftp.sanger.ac.uk/pub/project/pathogens/Phage/).

## Data Availability

All data produced in the present work are contained in the manuscript

## Acknowledgements

We thank the members of CHRF laboratory and environmental sampling team for valuable discussions and support. We would also like to thank Prof. Denise Monack and Dr. Ben Wang, Stanford University for providing laboratory strains used in this study.

## Supplementary Figures

**Supplementary Figure S1.**
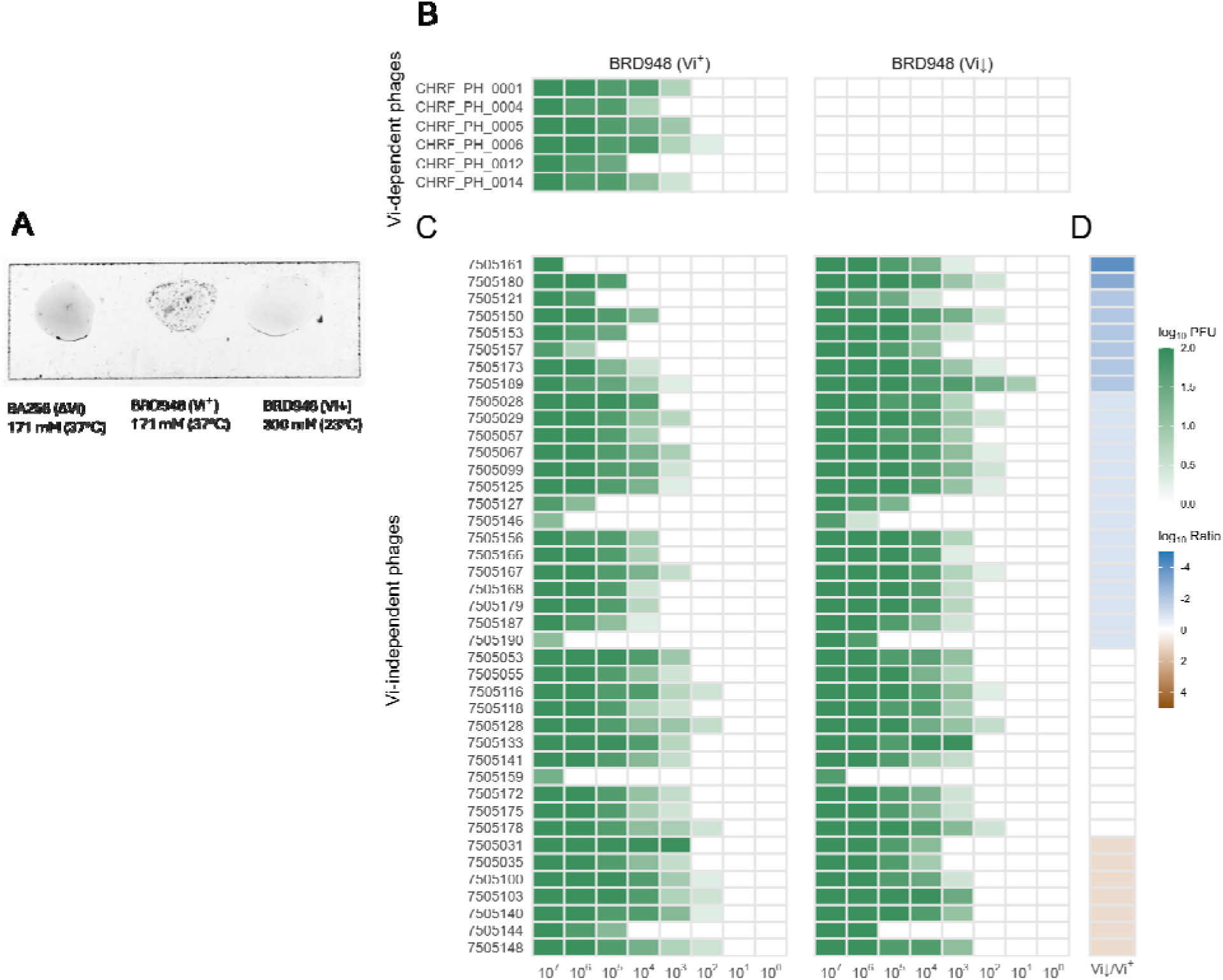
Capsule-dependent modulation of phage infectivity under Vi-suppressing conditions. (A) Vi antisera agglutination assay confirming capsule expression in BRD948 under standard conditions (Vi⁺, middle) and reduced capsule expression under Vi-suppressing conditions (ViS, right). (B, C) Infectivity heatmaps on Vi-expressing (Vi⁺) and Vi-suppressed (ViS) BRD948, shown as log₁₀(PFU + 1) for 6 Vi-dependent phages (B) and 41 Vi-independent phages (C). Vi⁺ and ViS cultures were grown in LB containing 171 mM NaCl at 37 °C and 300 mM NaCl at 23 °C, respectively. Column are 10-fold serial dilutions of lysate (10⁷–10⁰ PFU/mL); rows are individual phages. Darker green indicates higher infection efficiency; lighter colours indicate reduced or absent plaque formation. (D) Host-preference index. For each phage, the lowest PFU producing detectable plaques on ViS was compared with Vi⁺ and expressed as log₁₀(ViS/Vi⁺). Blue (−5 to −1), higher efficiency on ViS; white, similar efficiency on both; light brown (> 0), higher efficiency on Vi⁺. Generated in R (v4.5.2) with ggplot2.

**Supplementary Figure S2.**
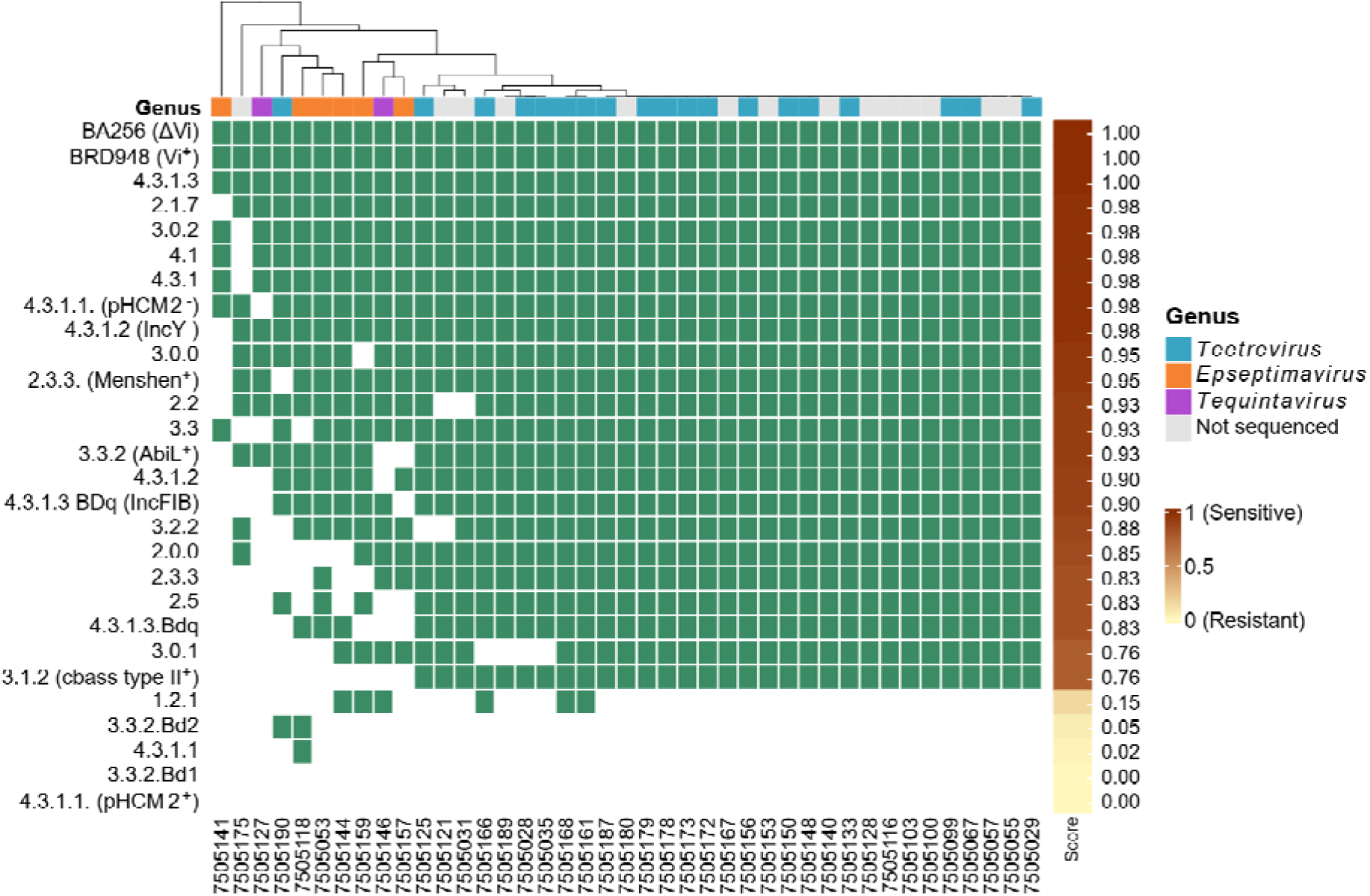
Infectivity spectrum of Vi-independent phages across circulating S. Typhi genotypes under Vi-expressing conditions. Infectivity of 41 Vi-independent phages against 26 circulating S. Typhi genotypes grown under Vi-expressing conditions (LB, 171 mM NaCl, 37 °C). Each phage was spotted at 10⁷ PFU/mL, and infectivity was scored by presence/absence of plaques. Green, detectable killing (sensitive); white, resistant. Rows are genotypes; columns are phages. The adjacent brown–yellow strip shows an infectivity score (0–1) relative to the BRD948 reference, which showed the highest susceptibility; scores near 1 resemble BRD948, scores near 0 indicate resistance. Hierarchical clustering highlights lineage-specific variation. The colored bar above the heatmap indicates phage genus. Generated in R (v4.5.2) with ComplexHeatmap; clustering used Jaccard distance (vegan).

**Supplementary Figure S3.**
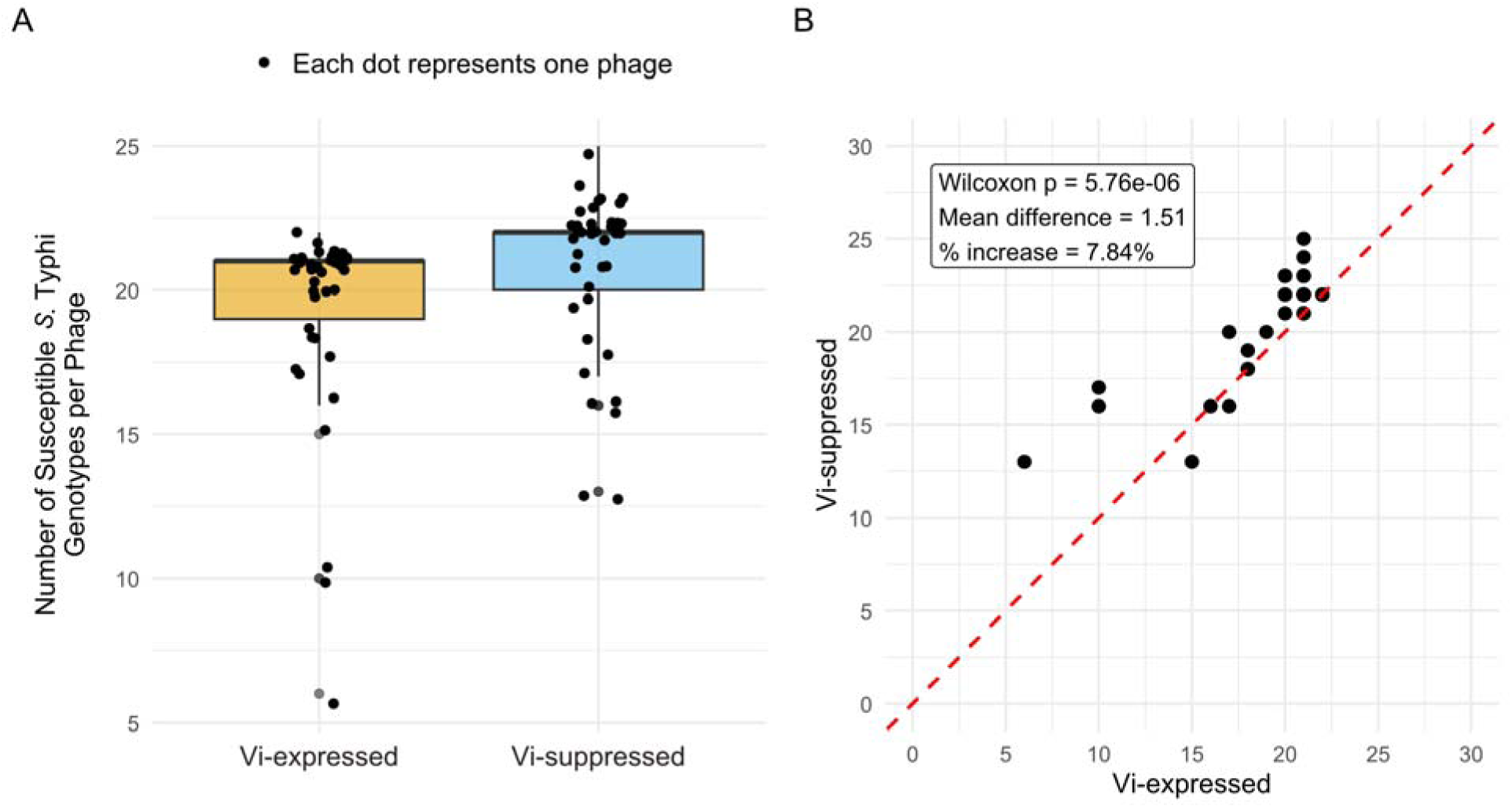
Quantitative comparison of Vi-independent phage infectivity under Vi-expressing and Vi-suppressed conditions. (A) Boxplot showing the number of S. Typhi genotypes susceptible to each phage under Vi-expressing (LB, 171 mM NaCl, 37 °C) and Vi-suppressed (LB, 300 mM NaCl, 23 °C) conditions; each dot represent one phage spotted at 10⁷ PFU/mL, and infectivity was scored by presence/absence of plaques. (B) Paired comparison of phage infectivity across conditions; each point represents a single phage, with the x-axis indicating susceptible genotype counts under Vi-expressing conditions and the y-axis indicating counts under Vi-suppressed conditions. Points above the dashed diagonal indicate increased infectivity under Vi-suppressed conditions. Conditions were compared by a two-sided Wilcoxon signed-rank test (p = 5.76 × 10⁻⁶). Generated in R (v4.5.2) with ggplot2.

**Supplementary Figure S4:**
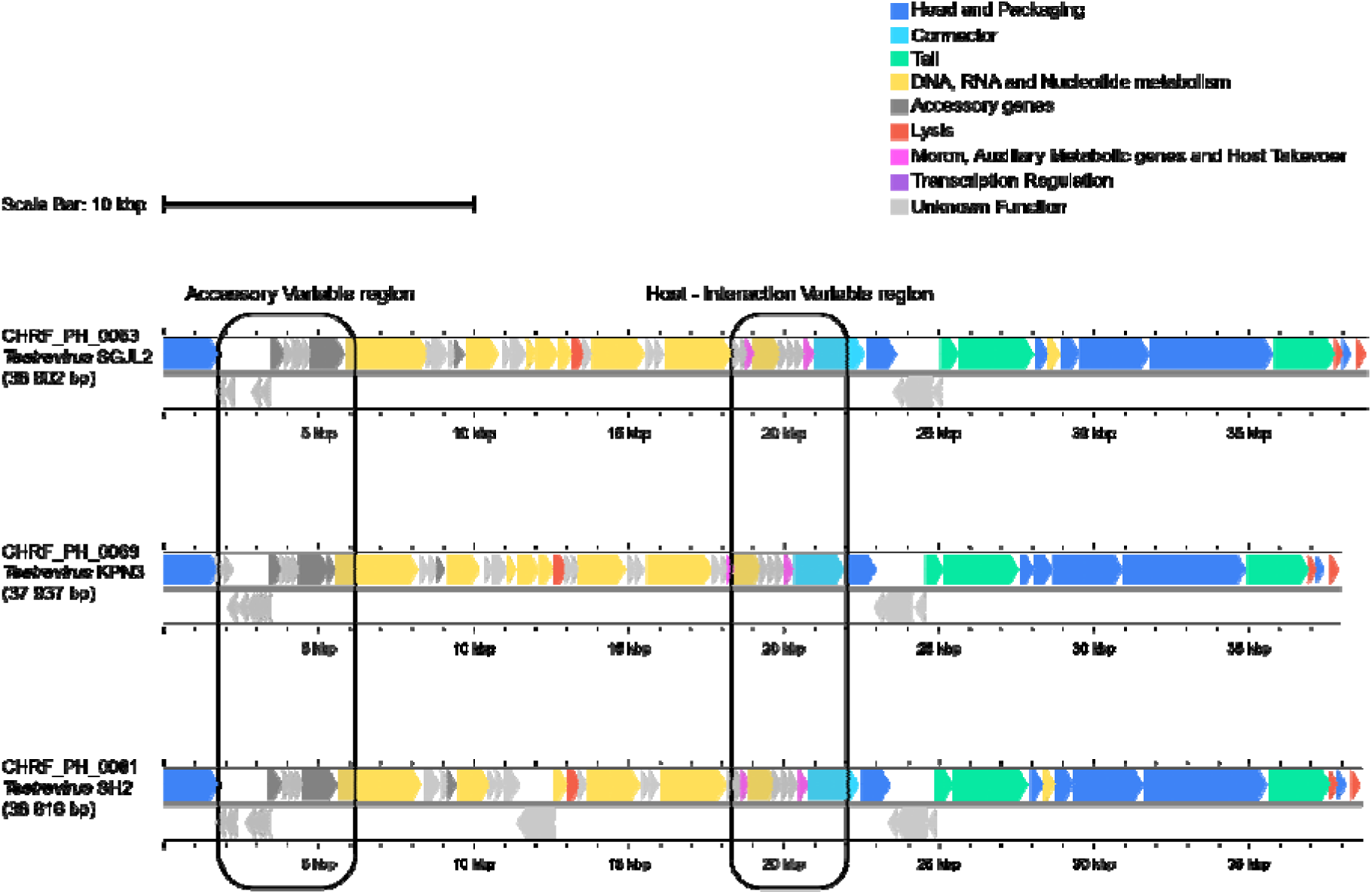
Genome map of the different sub-species of Teetreviru genus from the Autographiviridae family. Using the pharokka annotated GenBank files, genomic maps were generated in Proksee. Genes are colored according to their predicted functional categories. For consistency, the mapping is orientated with Terminase large subunit. Highlighted regions represent candidate modules identified through functional annotation, including genes involved with anti-defense or host interaction, along with hypothetical ORFs. The maps illustrate the overall distribution of genes and thei organization for functional roles across the genomes. Genome maps are drawn to scale, and a 10-kbp scale bar is provided.

**Supplementary Figure S5:**
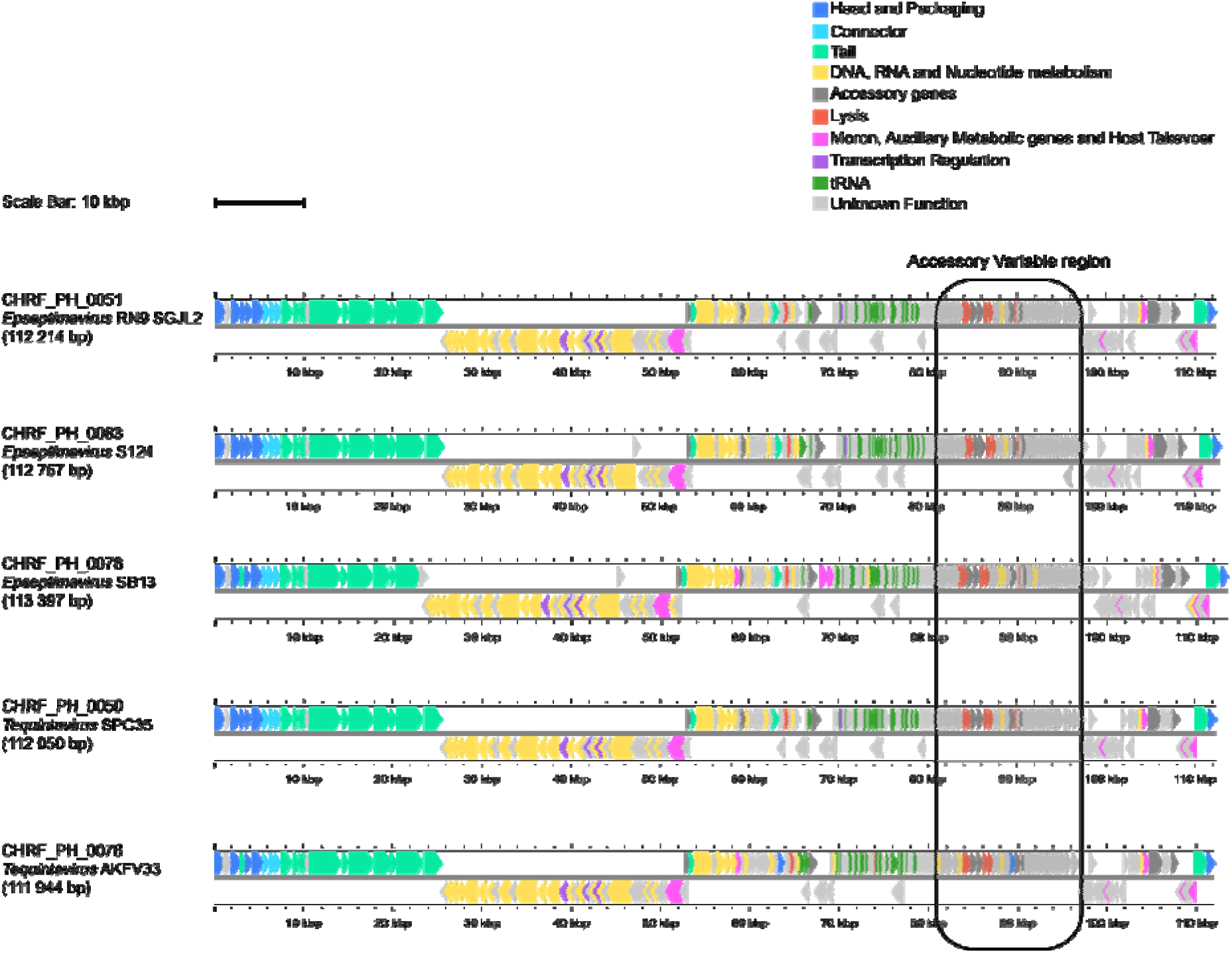
Genome map of the different sub-species of Epseptimavirus and Tequintavirus genus from the Demerecviridae family. Genome maps were generated in Proksee using Pharokka-annotated GenBank files. Genes were colored according to their predicted functional categories. The mapping is orientated with Terminase large subunit for consistency. Highlighted regions show candidate modules identified through functional annotation, including genes involved with anti-defense systems along with hypothetical ORFs. The maps illustrate the overall distribution of gene and their functional organization across the genomes. Genome maps are drawn to scale, and a 10-kbp scale bar is provided.

**Supplementary Table 1.** : Details of the 27 Vi-independent phages sequenced in this study

| NCBI<br>Accession<br>Number | Sample_ID | Phage_ID | Genome<br>Size,<br>bp<br>(ORF) | Coverage<br>(G+C<br>%content<br>) | Family/<br>Genus | Species<br>(Sub-species) |
| --- | --- | --- | --- | --- | --- | --- |
| SAMN5724888<br>3 | 7505028 | CHRF_PH_004<br>8 | 38 971<br>(53) | 3070x<br>(51) | *Autographiviridae*;<br>*Teetrevirus* | *Teetrevirus*<br>SGJL2<br>(*Salmonella*<br>phage ϕSG-JL2) |
| SAMN5724888<br>4 | 7505029 | CHRF_PH_004<br>9 | 38 415<br>(53) | 4235x<br>(51) | *Autographiviridae*;<br>*Teetrevirus* | *Teetrevirus*<br>SGJL2<br>(*Salmonella*<br>phage ϕSG-JL2) |
| SAMN5724888<br>5 | 7505146 | CHRF_PH_005<br>0 | 112<br>050<br>(203) | 1392x<br>(41) | *Demerecviridae*;<br>*Tequintavirus* | *Tequintavirus*<br>SPC35<br>(*Salmonella*<br>phage Spc35) |
| SAMN5724888<br>6 | 7505159 | CHRF_PH_005<br>1 | 112<br>214<br>(203) | 1098x<br>(40) | *Demerecviridae*;<br>*Epseptimavirus* | *Epseptimavirus*<br>s RN9<br>(*Salmonella*<br>phage<br>vB_STy-RN29) |
| SAMN5724888<br>7 | 7505187 | CHRF_PH_005<br>2 | 38 294<br>(54) | 3966x<br>(51) | *Autographiviridae*;<br>*Teetrevirus* | *Teetrevirus*<br>SGJL2<br>(*Salmonella*<br>phage ϕSG-JL2) |
| SAMN5724888<br>8 | 7505190 | CHRF_PH_005<br>3 | 38 802<br>(53) | 3507x<br>(51) | *Autographiviridae*;<br>*Teetrevirus* | *Teetrevirus*<br>SGJL2<br>(*Salmonella*<br>phage ϕSG-JL2) |
| SAMN5724888<br>9 | 7505035 | CHRF_PH_006<br>9 | 37 937<br>(56) | 2010x<br>(51) | *Autographiviridae*;<br>*Teetrevirus* | *Teetrevirus*<br>KPN3<br>(*Enterobacter*<br>phage KNP3) |
| SAMN5724889<br>0 | 7505053 | CHRF_PH_007<br>0 | 113<br>421<br>(207) | 793x<br>(40) | *Demerecviridae*;<br>*Epseptimavirus* | *Epseptimavirus*<br>s SB13<br>(*Salmonella*<br>phage<br>vB_SenS_SB1<br>3) |
| SAMN5724889<br>1 | 7505067 | CHRF_PH_007<br>1 | 38 428<br>(52) | 1421x<br>(51) | *Autographiviridae*;<br>*Teetrevirus* | *Teetrevirus*<br>SGJL2<br>(*Salmonella*<br>phage ϕSG- |
|  |  |  |  |  |  | JL2) |
| SAMN5724889<br>2 | 7505099 | CHRF_PH_007<br>2 | 39 019<br>(54) | 753x<br>(51) | *Autographiviridae;*<br>*Teetrevirus* | *Teetrevirus*<br>SGJL2<br>(*Salmonella*<br>phage φSG-JL2) |
| SAMN5724889<br>3 | 7505118 | CHRF_PH_007<br>4 | 11<br>1522<br>(210) | 641x<br>(40) | *Demerecviridae;*<br>*Epseptimavirus* | *Epseptimavirus*<br>SB13<br>(*Salmonella*<br>phage<br>vB_SenS_SB1<br>3) |
| SAMN5724889<br>4 | 7505125 | CHRF_PH_007<br>5 | 38 524<br>(51) | 1568x<br>(51) | *Autographiviridae;*<br>*Teetrevirus* | *Teetrevirus*<br>SGJL2<br>(*Salmonella*<br>phage φSG-JL2) |
| SAMN5724889<br>5 | 7505127 | CHRF_PH_007<br>6 | 111<br>944<br>(201) | 935x<br>(39) | *Demerecviridae;*<br>*Tequintavirus* | *Tequintavirus*<br>AKFV33<br>(*Escherichia*<br>phage<br>vB_EcoS_AKF<br>V33) |
| SAMN5724889<br>6 | 7505133 | CHRF_PH_007<br>7 | 39 281<br>(58) | 2545x<br>(51) | *Autographiviridae;*<br>*Teetrevirus* | *Teetrevirus*<br>KPN3<br>(*Enterobacter*<br>phage KNP3) |
| SAMN5724889<br>7 | 7505141 | CHRF_PH_007<br>8 | 113<br>397<br>(212) | 543x<br>(40) | *Demerecviridae;*<br>*Epseptimavirus* | *Epseptimavirus*<br>SB13<br>(*Salmonella*<br>phage<br>vB_SenS_SB1<br>3) |
| SAMN5724889<br>8 | 7505144 | CHRF_PH_007<br>9 | 115<br>413<br>(206) | 306x<br>(40) | *Demerecviridae;*<br>*Epseptimavirus* | *Epseptimavirus*<br>SB13<br>(*Salmonella*<br>phage<br>vB_SenS_SB1<br>3) |
| SAMN5724889<br>9 | 7505148 | CHRF_PH_008<br>0 | 38 304<br>(54) | 1844x<br>(51) | *Autographiviridae;*<br>*Teetrevirus* | *Teetrevirus*<br>SGJL2<br>(*Salmonella*<br>phage φSG-JL2) |
| SAMN5724890<br>0 | 7505150 | CHRF_PH_008<br>1 | 38 616<br>(53) | 1448x<br>(51) | *Autographiviridae;*<br>*Teetrevirus* | *Teetrevirus*<br>SH2<br>(*Citrobacter*<br>phage SH2) |
| SAMN5724890<br>1 | 7505156 | CHRF_PH_008<br>2 | 37 881<br>(52) | 625x<br>(51) | *Autographiviridae*;<br>*Teetrevirus* | *Teetrevirus*<br>SGJL2<br>(*Salmonella*<br>phage φSG-JL2) |
| SAMN5724890<br>2 | 7505157 | CHRF_PH_008<br>3 | 112<br>757<br>(204) | 98x<br>(40) | *Demereciviridae*;<br>*Epseptimavirus* | *Epseptimavirus*<br>S124<br>(*Salmonella*<br>phage S124) |
| SAMN5724890<br>3 | 7505161 | CHRF_PH_008<br>4 | 38 264<br>(54) | 664x<br>(51) | *Autographiviridae*;<br>*Teetrevirus* | *Teetrevirus*<br>SGJL2<br>(*Salmonella*<br>phage φSG-JL2) |
| SAMN5724890<br>4 | 7505166 | CHRF_PH_008<br>5 | 38 971<br>(53) | 1299x<br>(51) | *Autographiviridae*;<br>*Teetrevirus* | *Teetrevirus*<br>SGJL2<br>(*Salmonella*<br>phage φSG-JL2) |
| SAMN5724890<br>5 | 7505168 | CHRF_PH_008<br>6 | 38 294<br>(50) | 1602x<br>(51) | *Autographiviridae*;<br>*Teetrevirus* | *Teetrevirus*<br>KPN3<br>(*Enterobacter*<br>phage KNP3) |
| SAMN5724890<br>6 | 7505172 | CHRF_PH_008<br>7 | 38 667<br>(52) | 1031x<br>(51) | *Autographiviridae*;<br>*Teetrevirus* | *Teetrevirus*<br>KPN3<br>(*Enterobacter*<br>phage KNP3) |
| SAMN5724890<br>7 | 7505173 | CHRF_PH_008<br>8 | 38 537<br>(53) | 226x<br>(51) | *Autographiviridae*;<br>*Teetrevirus* | *Teetrevirus*<br>KPN3<br>(*Enterobacter*<br>phage KNP3) |
| SAMN5724890<br>8 | 7505178 | CHRF_PH_008<br>9 | 38 667<br>(52) | 880x<br>(51) | *Autographiviridae*;<br>*Teetrevirus* | *Teetrevirus*<br>KPN3<br>(*Enterobacter*<br>phage KNP3) |
| SAMN5724890<br>9 | 7505179 | CHRF_PH_009<br>0 | 38 264<br>(54) | 56x<br>(51) | *Autographiviridae*;<br>*Teetrevirus* | *Teetrevirus*<br>SGJL2<br>(*Salmonella*<br>phage φSG-JL2) |

